# Genes that cause severe liver disease in children also influence risk and severity of common liver conditions in adults

**DOI:** 10.1101/2025.05.09.25326407

**Authors:** JM Mushi, PJ Sharma, A Schofield, VL Chen, HJ Cordell, SP Davies, GL Gupte, GM Hirschfield, R Jeyaraj, DE Jones, GF Mells, YH Oo, RN Sandford, KA Siminovitch, J Xu, K Zhu, M Trauner, JP Mann

## Abstract

**Background and aims:** Rare, pathogenic variants can cause severe liver disease, requiring transplantation in childhood, but it is unclear how common variants in the same genes affect adults. Here, we aimed to establish population-level genetic evidence for whether ’monogenic’ diseases are associated with liver injury in adulthood.

**Methods:** We identified 99 genes where pathological mutations cause significant liver disease in children. For each, we used data from over 1.8 million adults to identify associations with biomarkers of liver injury. Observations were validated in multiple cohorts of adults with clinical liver disease and transcriptomics. Finally, we illustrated the importance of the JAG1-NOTCH pathway on the ductular reaction using immunohistochemistry.

**Results:** Most genes (56% (55/99)) had at least ’moderate’ evidence of association with liver-related traits at a population level. We identified 82 genome-wide (p<5x10^-8^) associations with markers of liver injury in 41% (41/99) of genes. Loss of function variants in these genes had a ten-fold greater effect on liver enzymes and well-established variants in *PNPLA3* had a three-fold greater effect. Variants in *ABCC2*, *ASL*, *BCS1L*, *HFE*, and *SERPINA1* were linked with presence of clinical liver disease in adults. Aggregated effects of 35 variants as polygenic risk score (PRS) was associated with 0.6% lower prevalence of MASLD between highest and lowest PRS groups. Transcriptional expression of 30% of genes was associated with severity of MASLD. Expression of JAG1-NOTCH2 pathway was associated with severity of PSC. JAG1 and NOTCH2 were expressed in injured bile ducts but not adjacent unaffected ducts.

**Conclusions:** Onset and severity of liver disease in adulthood is influenced by genes that also cause severe monogenic liver disease in children.

## Introduction

Germline variants with major functional impact cause rare, apparently monogenic diseases across all body systems. Such disorders (e.g. Alagille syndrome, Wilson disease, progressive familial intrahepatic cholestasis (PFIC)) are frequent indications for liver transplantation in children^1^. However, many conditions (e.g. Alagille syndrome) have wide clinical variation ^2^ and higher predicted genetic prevalence than observed clinical prevalence^3^. This may be due to incomplete penetrance, variable expressivity, and/or influences of environmental factors. Other disorders (e.g. progressive familial intrahepatic cholestasis) show close genotype-phenotype correlation, with more severe protein-truncating mutations requiring earlier transplantation^4^.

Large, population-scale sequencing studies have led to the identification of milder phenotypes of monogenic disorders across a range of conditions^5^. Such studies have demonstrated that variants in HFE (e.g. C282Y) and SERPINA1 confer additional risk towards chronic liver disease, consistent with an autosomal dominant method of inheritance^6,7^. However, it is not clear whether this principle extends to genes that cause ultra-rare monogenic liver diseases. Particularly as many disorders (e.g. *DCDC2*-associated neonatal sclerosing cholangitis) manifest a severe phenotype with biallelic mutations^8^ and are classically believed not to have a phenotype in the haploinsufficient state, though recent data has started to challenge this^9^.

Genetic testing for liver disease typically involves large, multigene panels. These often identify coding variants of uncertain significance for infants with neonatal cholestasis. Little is known about whether such variants confer risk later in life or have clinical relevance for the general population. In this study, we find evidence that ’monogenic’ liver disorders show mild phenotypes in adulthood.

## Materials and methods

### Identification of genes for inclusion

First, we identified a list of genes that are robustly associated with monogenic liver disease. Criteria for inclusion were: (1) the condition causes at least one of: moderate hepatic fibrosis, acute and/or recurrent liver failure, or primary liver cancer; (2) at least two separate case series of patients have been reported; (3) variants in a single gene are regarded as causal according to ACMG criteria^10^ review on Online Mendelian Inheritance in Man (OMIM)^11^. Genes were excluded if they were part of a generalised hereditary cancer syndrome (e.g. *TP53*), or if they were an indication for liver transplantation due to systemic effects without any evidence of clinically significant liver disease (e.g. *PKU*).

To identify genes, first, we searched PubMed and Google Scholar for: ("genetic" or "hereditary" or "monogenic") AND ("liver" or "biliary" or "fibrosis" or "hepatoblastoma" or "hepatocellular").

Relevant articles were reviewed to identify genes for inclusion. Next, we searched databases of genetic disorders (National Organisation for Rare Diseases (https://rarediseases.org/) and OMIM^11^) for the above liver-related terms. Lastly, we reviewed the Genomics England targeted sequencing panels (https://panelapp.genomicsengland.co.uk/) for Ductal plate malformation, Mitochondrial liver disease, including transient infantile liver failure, Neonatal cholestasis, and Polycystic liver disease.

### Gene-level association testing

Next, we determined the evidence for association between each of these 99 genes and liver-related traits using the HuGE calculator^12^ and GeneBass^13^ gene burden testing. The liver-related traits were: alanine transaminase (ALT), aspartate transaminase (AST), alkaline phosphatase (ALP), gamma-glutamyl transferase (GGT), total and conjugated bilirubin, cirrhosis, cholecystitis, and cholangitis. For each gene, we obtained HuGE scores for liver-related traits from the Common Metabolic Diseases Knowledge Portal (CMDKP, cmdkp.org, accessed February 2025). HuGE scores are calculated using the effect of both thousands of common and rare variants in (or near) each gene from multiple GWAS with sample sizes up to 1 million individuals, as described elsewhere in detail^12^. Evidence is graded from ’anecdotal’ to ’compelling’, after adjustment for multiple testing.

GeneBass is a curated database of results from UK BioBank whole exome sequencing and methods used for burden testing are described in detail elsewhere^13^. In short, the effects of multiple variants within a single gene are added together, based upon their predicted impact on protein function. Variants are classified into those predicted to cause loss of function (pLoF), missense mutations, and those without effect on amino acid sequence (synonymous). We extracted pLoF, missense, and synonymous gene burden testing (using SKAT-O p-values) and adjusted for multiple testing using Benjamini-Hochberg method.

### Variant-trait associations

We then identified variants in (or near (±50kb)) each of the 99 candidate genes that were associated with liver-related traits at genome-wide significance (p<5x10^-8^). To do this, we obtained variant-trait associations from CMDKP, GeneBass, and OpenTargets Genetics^14^. UCSC LiftOver Tool^15^ was used to convert from GRCh37 to GRCh38. We filtered for liver-related traits and genome-wide significant variants. Where duplicates were identified (from different data sources), the association with the smallest p-value was kept. To adjust for multiple testing, we only kept variants with p<5x10^-8^.

### Variant annotation

We annotated variants with functional predictions using Ensembl’s variant effect predictor^16^. LocusZoom was used to illustrate locus-trait associations^17^. Variants were further annotated with expression-trait quantitative loci (eQTL) in liver tissue using data from Genotype-Tissue Expression (GTEx) Project via the EBI eQTL catalogue^18^.

To determine whether causal variants were shared across traits (e.g. ALT and GGT), we performed colocalization analysis using summary statistics from the largest available GWAS of liver enzymes^19^ (as full bottom-line association results were not available from CMDKP).

Summary statistics were standardised using MungeSumstats^20^ and converted into variant call format using gwas2vcf^21^ for analysis using gwasglue. Evidence of colocalization due to a single causal variant was defined by PP.H_4_ ≥0.8^22^.

We manually reviewed evidence from Open Targets Genetics^14^ and FORGEdb^23^ for every variant to determine whether the look-up gene was likely to be causal. For variants to be included as likely linked to the look-up gene required, at least one of: coding or intronic within the gene of interest, significant eQTL, or in strong LD (R^2^>0.8) with another variant that fulfilled the above criteria.

To determine the number of unique variants, we subsequently performed clumping using 1000 genomes phase 3^24^ reference data with PLINK2^25^ --clump-p1 1e-5 --clump-r2 0.05 --clump-kb 250. LeadSNPs were then taken forwards for subsequent validation.

### Clinical liver cohort validation

Next, we replicated associations in separate genome-wide association studies of patients with clinically significant common liver diseases. We obtained summary statistics from six studies: primary sclerosing cholangitis (PSC)^26^, primary biliary cholangitis (PBC)^27^, metabolic-dysfunction associated steatotic liver disease (MASLD)^28^, and all-cause cirrhosis^6,29,30^. None of these studies were included in the discovery searches. Replication required false-discovery rate (FDR (Benjamini-Hochberg)) adjusted p-FDR < 0.05 in at least one cohort with clinical liver disease.

### All of Us validation and polygenic risk score

All of Us is a population-level dataset of approximately 350,000 individuals from the United States of America. Inclusion criteria and genotyping of the All of Us cohort are described in detail elsewhere^31^. All of Us data was accessed using Research Project ID 69402. Genetic analyses were restricted to the independent GWAS-significant variants identified above. 35 out of 82 variants were present in the dataset. Individuals were included in the current analysis if there was available data on age, sex, liver enzymes, BMI, and presence or absence of chronic liver disease. ICD diagnostic codes were used to identify cases of chronic liver disease; full details of diagnostic codes used are available in code. Diagnoses of MASLD were established using codes K75.8, K76.0, and K70.1. Genomic analyses were conducted using Hail^32^.

Individual variant-trait analyses were adjusted for age, sex, and the first 9 principal components (PC) of genetic ancestry. Bonferroni-adjusted p-value of .05/35= .0014 was used for significance.

We then constructed an unweighted polygenic risk score (PRS) through aggregation of number of alternate alleles (as identified in the original variant discovery above) for each individual. The presence or absence of clinical liver disease was tested against PRS using multivariable logistic regression adjusted for age, sex, and body mass index (BMI).

### Gene expression in MASLD

We then assessed whether expression of the candidate genes contribute to severity adult liver disease. Transcriptional expression of 99 genes was assessed in GSE135251^33^, which contains RNA sequencing data from 216 individuals across the spectrum of MASLD (control to F4). Raw counts were filtered for low expressed genes then differential gene expression was performed between patients with MASLD and no fibrosis (MASL F0) and steatohepatitis with cirrhosis (MASH F4) with Benjamini–Hochberg adjusted p-values <0.05 as significant. 94 out of 99 genes were available for analysis in the dataset. Violin plots demonstrate normalised counts. In addition, we performed one-way ANOVAs to look for any variation between groups in candidate genes, with Bonferroni-adjusted p-values (p.adj) <0.05 as significant.

### JAG1 transcript expression in PSC and MASLD

Given the strong signal for JAG1 in the above analyses we aimed to validate the transcriptional results in additional cohorts of PSC and MASLD. Patients with PSC who were recruited to the phase 2b trial of simtuzumab (NCT01672853)^34^ had RNA extracted from FFPE baseline liver biopsies. RNA with DV200 >10% underwent cDNA synthesis using Illumina TruSeq RNA followed by paired-end 150bp read sequencing on Illumina NovaSeq 6000. Only data that passed quality control (Qscore, AT/GC distribution, and error rate distribution) were subject to further analysis. FASTQ files had adapter sequences removed using cutadapt^35^ and were then aligned to GRCh38 human reference genome using STAR^36^. Samtools^37^ and featureCounts were used for sorting and counting. Log_2_ transcripts per million (TPM) were used to compare JAG1, NOTCH1, HEYL, and HES expression between patients with mild (Ishak 0-2), moderate (Ishak 3-4), and severe fibrosis (Ishak 5-6) using Kruskal-Wallis test, with p-values adjusted for multiple testing. These results were replicated using the SteatoSITE MASLD transcriptomic dataset^38^ available from https://shiny.igc.ed.ac.uk/SteatoSITE_gene_explorer/.

### Immunohistochemistry (IHC)

IHC was performed on liver tissue from explants obtained at the time of transplant from patients with PSC, PBC, alcohol-related liver disease (ARLD), and MASLD, as well as healthy donor liver. The Queen Elizabeth Hospital in Birmingham is a major liver transplant centre performing more than 300 transplants per year. All samples used in this study were obtained as part of the routine biobanking of explanted (typically cirrhotic) liver tissue taken from transplants^39^. All liver tissue was obtained following informed consent, under the "Liver Inflammation" study (Research Ethics Committee reference: 18/WA/0214; IRAS reference: 223072). Fresh tissue was formalin-fixed and paraffin-embedded (FFPE). FFPE sections were dewaxed in xylene then rehydrated in graded alcohols before heat antigen retrieval using Tris (Vector, H-3301) for 30 minutes.

Endogenous peroxidase activity was blocked using BLOXALL (Vector, SP-6000-100) for 15 minutes at room temperature. Non-specific binding was blocked for 20 minutes at room temperature (Merck Millipore, 20773). Primary antibodies were incubated overnight at 4°C. Primary antibodies used were: JAG1 (Cell Signalling, D4Y1R, at 1/200), NOTCH2 (Cell Signalling, D76A6, at 1/50), and GBA1 (Abcam, EPR5142, at 1/200). Secondary antibodies (MP-7451 and MP-7452, Vector) were incubated for 1 hour at room temperature followed by developing with ImmPACT DAB (Vector, SK-4105) for 5 minutes and counter-stained with Mayers hematoxylin.

### Immunofluorescence

Multiplex immunofluorescence was also performed on human liver FFPE samples. Antigen retrieval and blockade was performed as above. Each of four primary antibodies (JAG1, NOTCH2, CK19 (Abcam, Ab220193, at 1/1000), and ACTA2 (L-smooth muscle actin, Abcam, ab5694)) were incubated at room temperature for 1 hour. This was followed by 30 minutes of HRP-conjugated secondary antibody, as above, then 10-minutes development with tyramide signal amplification reagents: fluorescein (Akoya, NEL741001KT), Cy3 (Akoya, NEL744001KT), AZDye594 (CCT-1542), Cy5 (Akoya, NEL745001KT). Antibodies were stripped using Vectaplex (Vector, VRK-1000) for 15 minutes + 15 minutes at room temperature. Then blocking was repeated before the next primary antibody. Autofluorescence was quenched with TruView (Vector, SP-8400-15) and slides mounted with Vectashield with DAPI (Vector, H-1800). Images were acquired on a Zeiss LSM880 Confocal Microscope.

### Statistical analysis

Integration of datasets was performed using R base functions and tidyverse. Genome-wide significance (p<5x10^-8^) was used for initial identification of variant-trait associations with p<0.05 adjusted for multiple testing used throughout otherwise. All analyses were performed using R. Code is available from https://github.com/jmann01/MonogenicLiver.

## Results

### Monogenic liver disease genes demonstrate population-level associations

We identified 99 monogenic disorders that cause significant liver disease (Fig 1A & SupTab 1). Of these, 56% (55/99) had at least ’moderate’ evidence of association with population-level liver-related traits using the HuGE calculator (Fig 1B) whilst 17% (17/99) were associated with at least two traits with ’very-strong’ or ‘compelling’ evidence (Fig 1D & SupTab 2). This replicated well-established associations between *SERPINA1* (which causes alpha-1-antitrypsin deficiency) and *HFE* (which causes haemochromatosis) on ALT levels (Fig 1C). Genes that cause ductal plate malformations (e.g. *PKD1* and *PKHD1*) clustered together with genes that cause progressive familial intrahepatic cholestasis (e.g. *NR1H4*) due to associations with serum liver enzymes.

**Fig 1.**
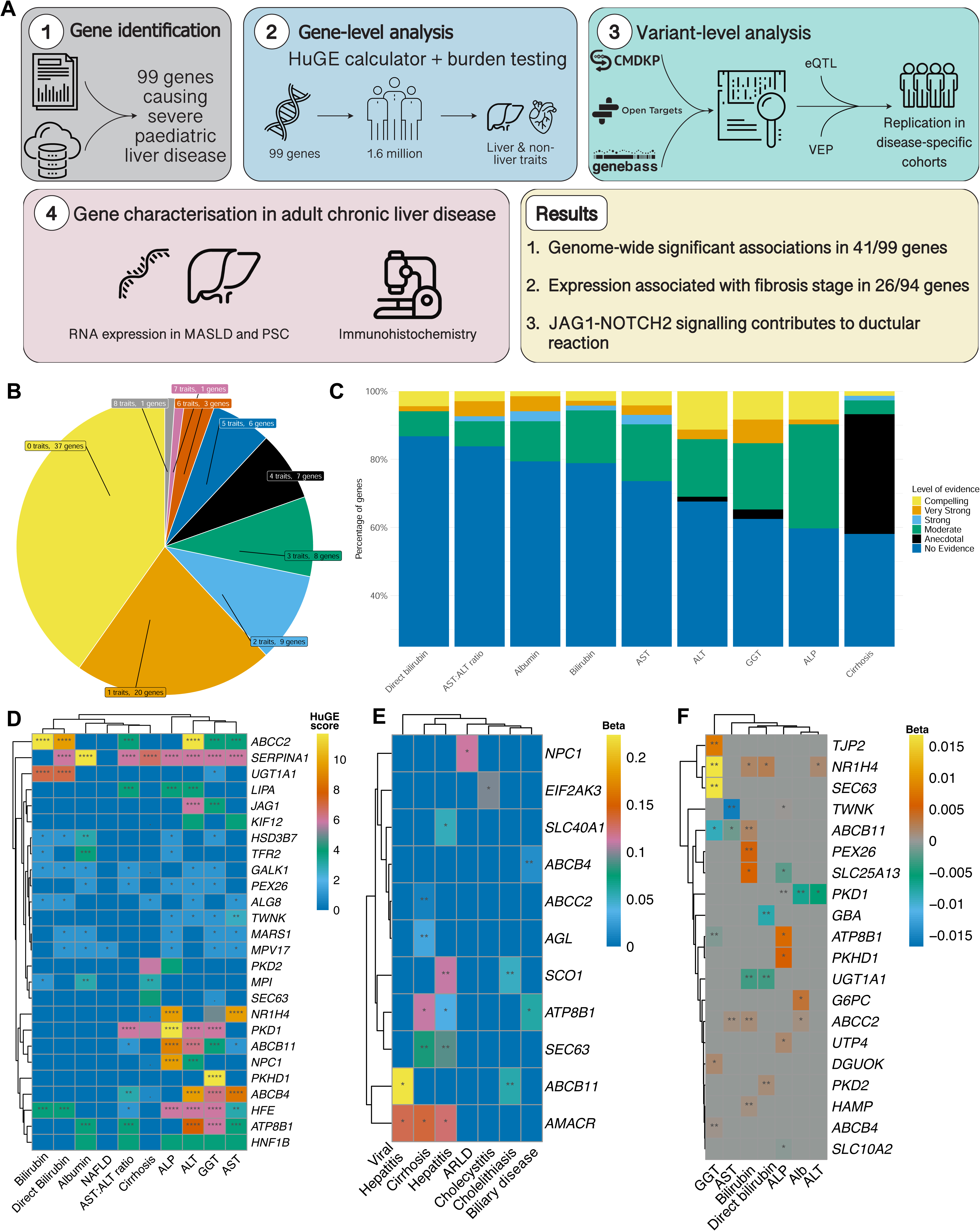
Genes that cause monogenic liver disease are associated with liver-related traits at a population level. (A) Summary of the study design. (B) Proportion of genes with at least ’moderate’ evidence for liver-related traits based on HuGE score (Human Genetic Evidence score from the Knowledge Portal Network). (C) Proportion of genes with each level of evidence for liver-related traits based on HuGE score. (D) Heatmap showing standard deviations of HuGE score (normalised by trait) and level of evidence for liver-related traits: * = Moderate, ** = Strong, *** = Very strong, **** = Compelling. (E-F) Heatmap results from gene burden testing on GeneBass showing standard deviations of beta for association for binary (E) and continuous (F) traits, annotated with pFDR: * ≤.05, ** ≤.01, *** ≤.001, *** ≤.0001. ALP, alkaline phosphatase; ALT, alanine aminotransferase; ARLD, alcohol-related liver disease; AST, aspartate aminotransferase; GGT, gamma-glutamyl transferase; MASLD, metabolic-dysfunction-associated steatotic liver disease.

These findings were replicated using gene burden testing in the UK BioBank: 44% (44/99) of genes demonstrated at least one association (at p-FDR<0.05, Fig1E-F & SupTab 3). Similar patterns of gene-trait associations were demonstrated across pLoF, missense, and synonymous burden testing.

### Common variants in monogenic disorder genes show genome-wide level associations with liver enzymes

We identified 82 independent variants associated with liver-related traits in 41% (41/99) of genes that cause ’monogenic’ liver disease (Table 1 and SupTab 4). 84% (69/82) were common variants (>1% effect allele frequency).

**Table 1.**
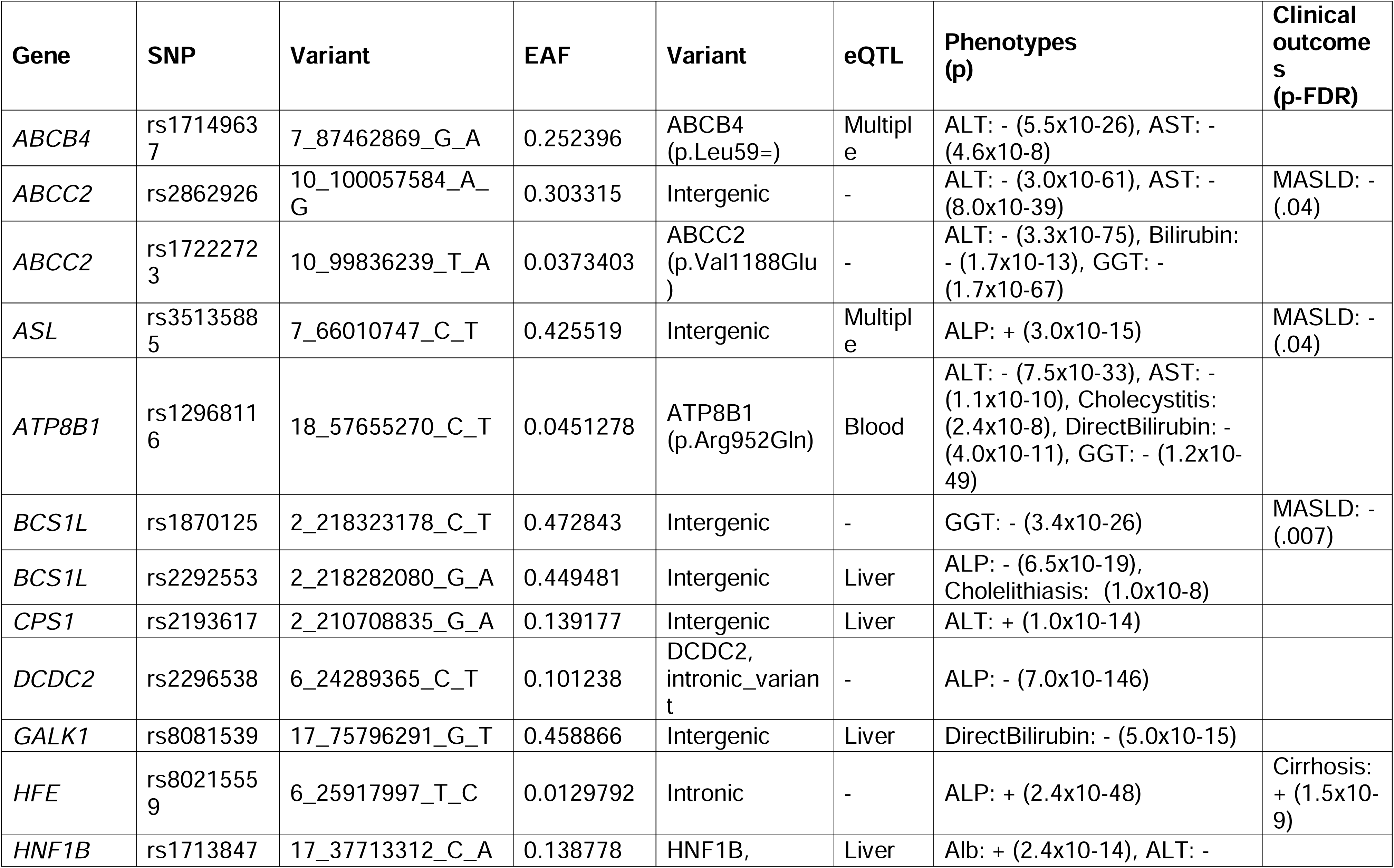
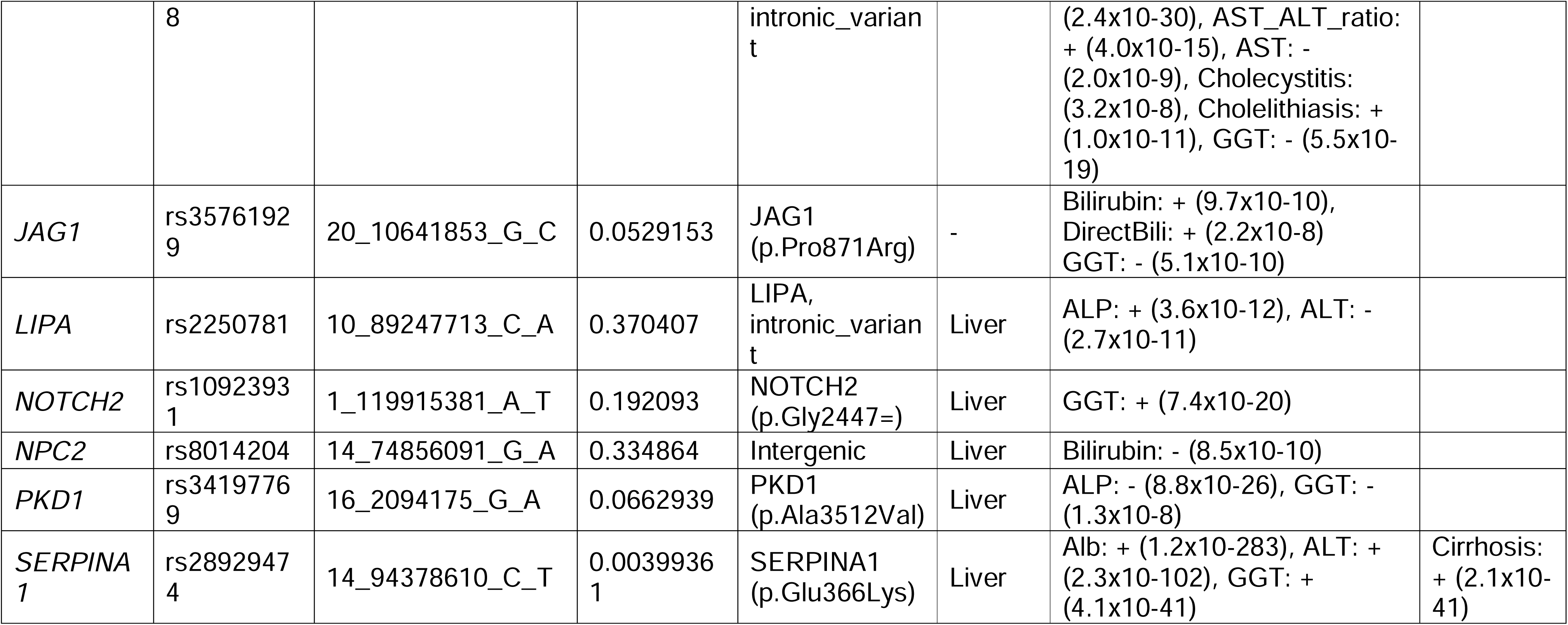
Variant-trait associations in genes that also cause monogenic liver disease. Variants are annotated with: predicted protein functional effect (from Ensembl’s Variant Effect Predictor), liver tissue eQTL data from GTEx, effect allele frequency (EAF) from gnomAD, and results from replication across six cohorts of patients with clinical liver disease.

Genome-wide significant associations were observed across all studied traits (Fig 2A). This replicated well-established associations between variants in *UGT1A1* (which causes Crigler-Najjar syndrome, as well as the more benign Gilbert’s syndrome) and bilirubin. Where genes were associated with more than one phenotype (e.g. *ABCB4* was associated with GGT and ALT), colocalization analyses demonstrated that this was likely due to a single causal variant (Fig 2B-E & SupTab 5). For example, a single locus within *HNF1B* was associated with GGT, ALP, and albumin (Fig 2C).

**Fig 2.**
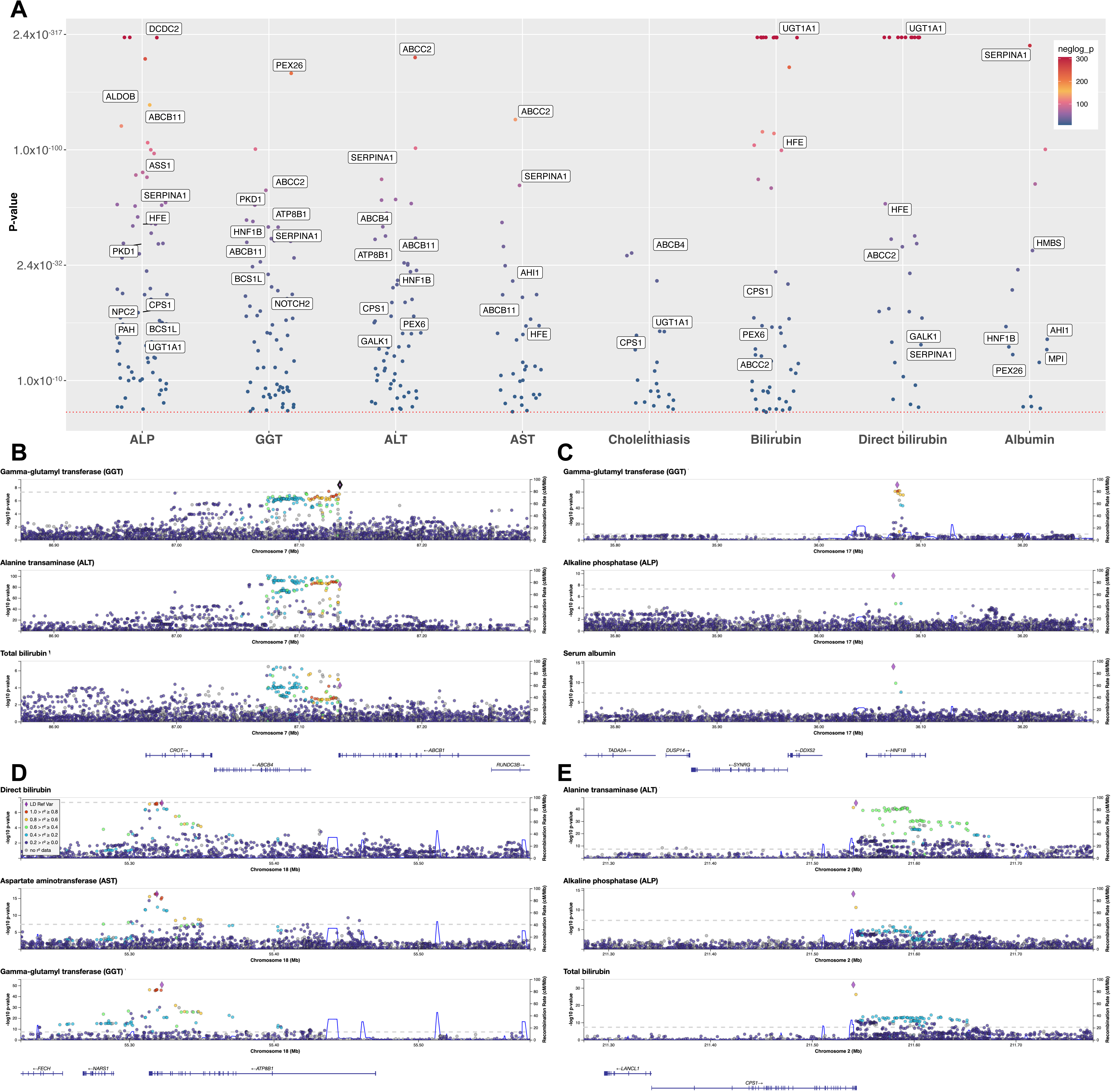
Common variants in genes that cause monogenic disease are associated with liver-related traits. Dot plot showing individual variants P-values for liver-related traits, with annotation of a selection of top genes (A). Y-axis is log_10_(-log_10_(p-values)) scaled. LocusZoom plots from Common Metabolic Disease Knowledge Portal Data demonstrating associations for *ABCB4* (B), *HNF1B* (C), *ATP8B1* (D), and *CPS1* (E).

Variants in genes that cause bile acid disorders, ductal plate malformations, and intrahepatic cholestasis predominantly had associations with indices of biliary disease (i.e. GGT, bilirubin, cholelithiasis, SupTab 4). For example, rs17855121T>C near *HSD3B7* (which causes 3β-hydroxy-Δ5-C27-steroid dehydrogenase deficiency, OMIM #607765) was associated with lower bilirubin levels (β (SE) = -0.015 (0.003), p=3.0x10^-8^) and rs16961277A>G in SLC10A2 (which causes Primary bile acid malabsorption 1, OMIM #613291) was associated with cholelithiasis and cholecystitis (β (SE) = -0.08 (0.012), p=1.4x10-^14^).

24% (20/82) were protein-coding variants in the gene of interest. This included missense variants in *PKD1*: p.Pro36His, p.Arg1340Trp, p.Thr2250Met, p.Arg2765Cys, all of which affect 1 in 500-700 individuals and therefore are too common to cause autosomal dominant polycystic kidney disease in isolation. All these variants are associated with GGT and/or ALP at genome-wide significance (all p≤2.9x10^-9^ ,SupTab 4).

Three missense variants (p.Val417Ile, p.Gly921Ser, p.Val1188Glu) in *ABCC2* (which causes Dubin-Johnson syndrome) were associated with multiple indices of liver injury: ALT, AST, GGT, and (conjugated) bilirubin (all p≤3.6x10^-9^).

For the non-coding variants, 44% (27/62) had a significant eQTL for the gene of interest (SupTab 4). For example, an intronic variant in *HNF1B* (which causes the dominant disorder Renal cysts and diabetes syndrome, OMIM #137920) was associated with liver-specific eQTL for *HNF1B* (p=6.0x10^-3^) and genome-wide associations in albumin, ALT, AST, GGT, cholelithiasis, and cholecystitis (all p≤3.2x10^-8^).

Within genes there is a dose-effect response, with more severe loss-of-function variants having greater effect sizes. Data from rare variant analysis on the Common Metabolic Disease Knowledge Portal demonstrates that, pooling 53 loss of function (LoF) variants in *ABCB4* are associated with higher ALT with nearly 10-fold greater effect size compared to pooling of 511 missense variants (β = 0.026 LoF versus β = 0.0024 for missense). Similarly, 100 LoF variants in ABCC2 on direct bilirubin had about 10 times the effect size compared to 647 missense variants (β = 0.011 LoF versus β = 0.0019 for missense).

The magnitude of effect of these variants is smaller than well-established common variants that influence liver disease. For example, rs41318029:G>A in *ABCC2* was associated with GGT with β = 0.06, whilst, a common variant in *GGT1* (rs3859862:A>G, effect allele frequency = 0.42) has more than three times the effect size with β = 0.21. Similarly, rs1047891:C>A in *CPS1* is associated with higher ALT with β = 0.04, whereas the well-established variant in *PNPLA3* rs738409:C>G has again three times the effect size at β = 0.15.

Overall, this demonstrates that common variants in genes that cause monogenic liver disease are associated with indices of liver injury.

### Variants in five genes validated with clinical liver disease outcomes

Next, we sought to validate individual variants in independent cohorts of adults with clinically significant liver disease.

Using six GWAS in patients with liver disease we established that variants in five genes (*ABCC2*, *ASL*, *BCS1L*, *HFE*, and *SERPINA1*) were associated with at least one clinical liver disease (SupTab 6 & Table 1). This included the well-established all-cause cirrhosis risk variants in *SERPINA1* and *HFE*^29,30^.

### Cumulative burden of variants in monogenic disorder genes affect risk of chronic liver disease

We extended these observations by replication in the All of Us cohort in nearly 350,00 individuals. 35 out of 82 independent variants were present in the All of Us dataset and we replicated variant-trait association across 57% (20/35) variants (SupTab 7).

Then we created an unweighted polygenic risk score (PRS) using the number of effect alleles across all 35 variants. Consistent with the effect direction observed in the original discovery, higher PRS quartile was associated with lower levels of ALP, ALT, and AST, but higher total and conjugated bilirubin (Table 2). The effect sizes are small: from highest to lowest quartile there is 0.1 IU/L difference in ALT and ALP. However this translated into a significant negative association with diagnosis of MASLD (β (SE) = -0.0116 (0.0031), p=1.7x10^-4^) or any chronic liver disease (β (SE) = -0.0050 (0.0022), p=.02) after adjusting for age, sex, and BMI (Figure 3 and SupTab8). Between PRS

**Fig 3.**
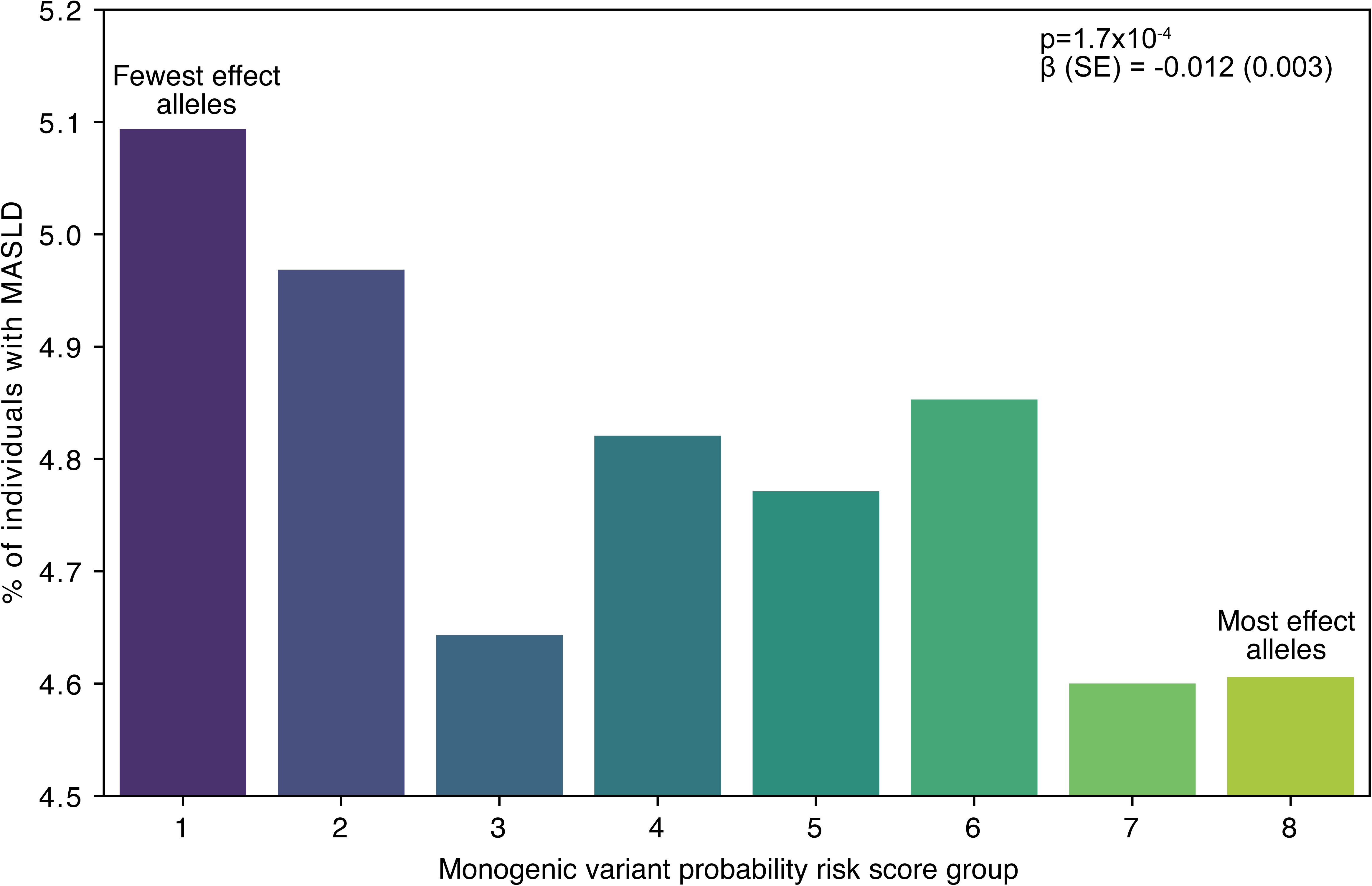
Prevalence of metabolic dysfunction-associated steatotic liver disease (MASLD) in the All of Us study, stratified by monogenic polygenic risk score (PRS). PRS was calculated as the sum of alternate alleles (without weighting) for all 35 variants present in the dataset. P-value was derived using multivariable logistic regression in n=346,332 individuals dataset for the presence of MASLD against (PRS), adjusted for age and sex.

**Table 2.** Monogenic gene polygenic risk score (PRS) is associated with liver enzymes and bilirubin levels. An unweighted PRS comprised of 35 variants in genes that cause monogenic liver disease was calculated in 291,758 individuals from the All of Us study. ANNOVA was used to compare the differences in liver enzymes between quartiles (Q1-Q4) of the PRS.

|  |  | Unweighted monogenic gene PRS quartile |  |  |  |  |
| --- | --- | --- | --- | --- | --- | --- |
| Trait | All | Q1 | Q2 | Q3 | Q4 | P-value |
| N<br>(% male) | 291758<br>(40%) | 88329<br>(39%) | 81193<br>(40%) | 69388<br>(40%) | 52848<br>(40.0%) | - |
| ALP (IU/L) | 76.2<br>(21.6) | 76.4 (21.8) | 76.3 (21.6) | 76.0 (21.5) | 75.8 (21.3) | 3.23E-05 |
| ALT (IU/L) | 23.3<br>(10.0) | 23.5 (10.1) | 23.3 (9.9) | 23.1 (9.9) | 23.2 (9.9) | 6.29E-09 |
| AST (IU/L) | 22.9 (7.0) | 23.0 (7.1) | 22.8 (7.0) | 22.8 (7.0) | 22.8 (7.0) | 0.00047621 |
| Total<br>bilirubin<br>(mg/dL) | 0.518<br>(0.21) | 0.504 (0.20) | 0.513 (0.21) | 0.523 (0.21) | 0.542 (0.22) | 1.11E-151 |
| Conjugated<br>bilirubin<br>(mg/dL) | 0.145<br>(0.07) | 0.145 (0.07) | 0.147 (0.07) | 0.148 (0.07) | 0.151 (0.07) | 3.08E-18 |

### Monogenic disorder genes show expression changes in MASLD and PSC progression

Given that most variants identified in our analysis were non-coding, we hypothesised that they may be acting through gene dosage. Therefore, we determined whether expression of these genes change with severity of liver disease using n=216 individuals with MASLD. 38% of genes (26/94) demonstrated variance with severity of MASLD (Fig 4 & SupTab 9). Specifically, 30% (28/94) showed differential expression between patients with MASL and MASH cirrhosis. This included upregulation and downregulation of genes across multiple different pathways (e.g. *DCDC2*, *HNF1B*, *HJV*). The top differentially expressed gene was *JAG1* (Fig 4E, log2FC 1.6, p.adj = 2.9x10^-13^). This finding was replicated in primary sclerosing cholangitis *JAG1* (Fig 4I and SupFig 1) and an additional MASLD cohort (Fig 4J).

**Fig 4.**
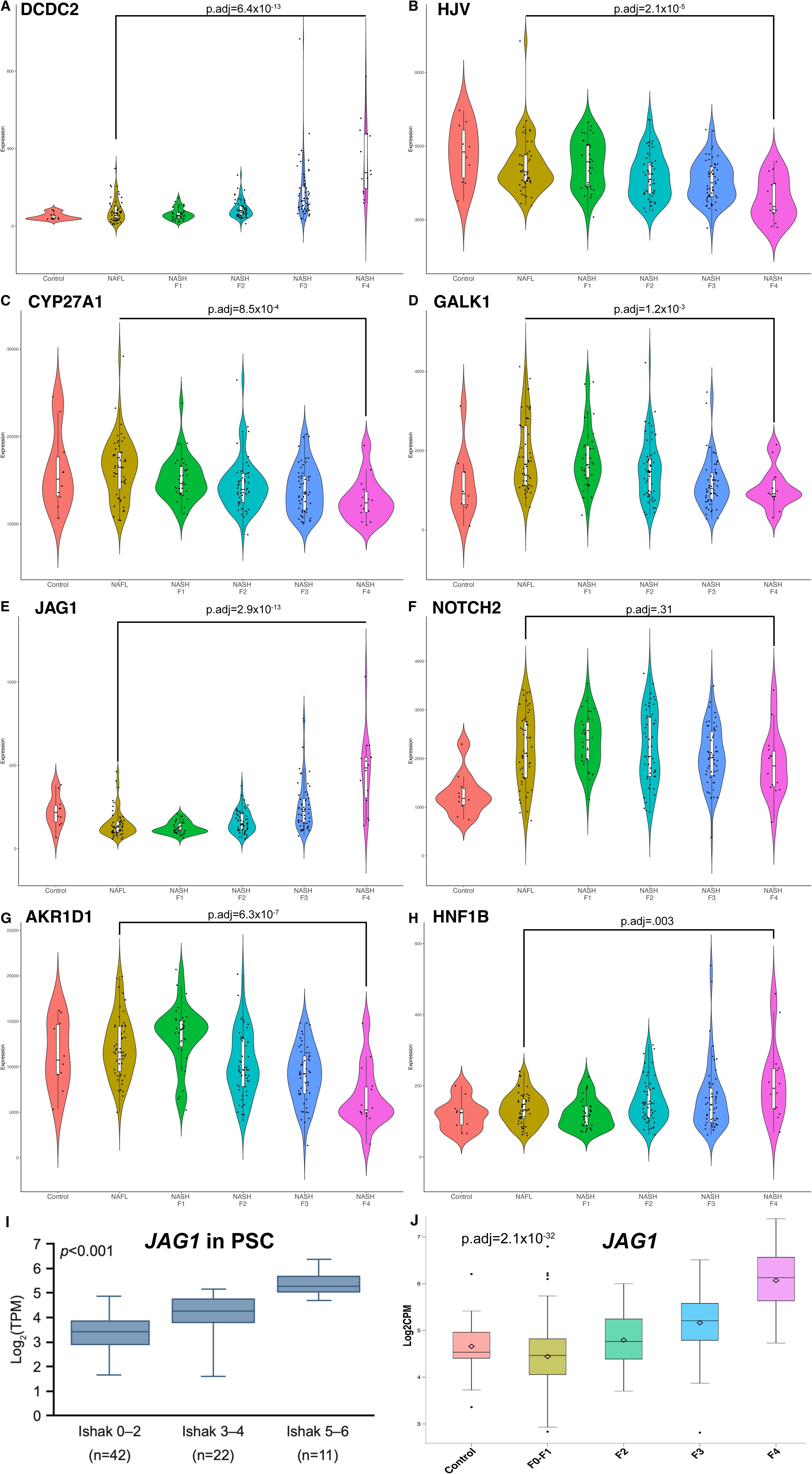
Expression of genes that cause monogenic disease is associated with severity of MASLD. Violin plots showing normalised gene expression across the spectrum of MASLD (n=216) from GSE135251 for *DCDC2* (A), *HJV* (B), *CYP27A1* (C), *GALK1* (D), *JAG1* (E), *NOTCH2* (F), *AKR1D1* (G), and *HNF1B* (H). p.adj represent differential gene expression between MASL and MASH F4 groups. Association of *JAG1* transcripts with fibrosis stage in PSC^34^ (E) and MASLD^38^ (F).

### JAG1-NOTCH signalling localises to ductular reaction in chronic liver disease

Our analyses found multiple variants in *JAG1* associated with indices of liver injury in adults as well as transcriptionally associated with stage of liver disease. Therefore, we next performed immunohistochemistry to visualise how JAG1-NOTCH signalling is perturbed in chronic liver disease. JAG1 is expressed in arterial structures in healthy adult liver (Fig 5A). However, it becomes strongly expressed in regenerative biliary ductules as part of the ductular reaction, most evident in cholangiopathies but also in ARLD, as previously reported^40^. NOTCH2 was not readily detectable in healthy liver but widely expressed throughout fibrotic areas and reactive ductules in cirrhosis (Fig 5B). Using multiplex immunohistochemistry, we found that JAG1 and NOTCH2 are co-expressed on injured biliary epithelial cells (CK19^+^LSMA^+^) in PSC and PBC but not on adjacent unaffected ducts (Fig 5C). In addition, NOTCH2 was expressed on a subset of LSMA^+^ fibroblasts within fibrotic regions.

**Fig 5.**
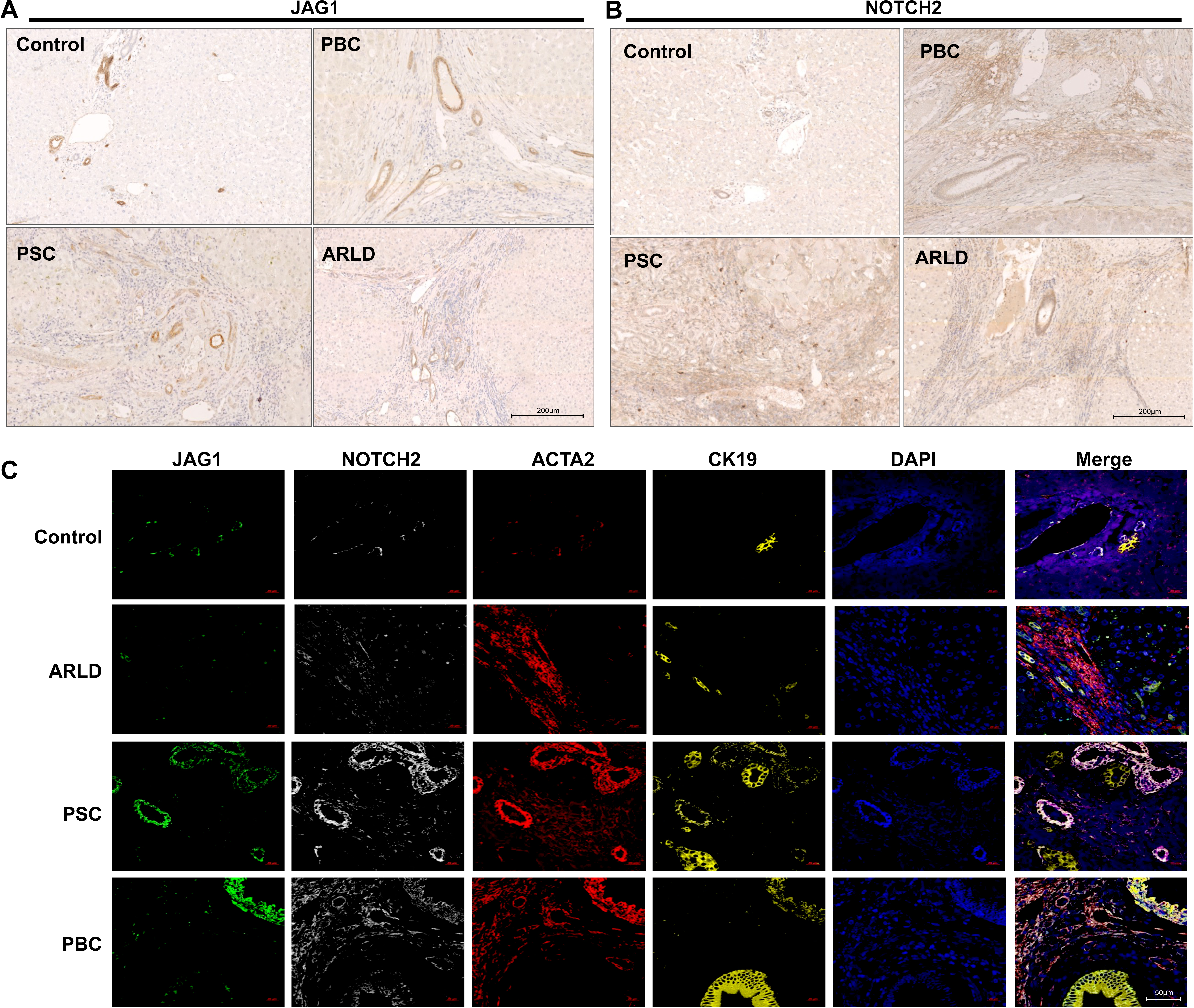
JAG1-NOTCH expression on injured bile ducts affects severity of PSC. Expression of JAG1 and NOTCH2 in normal liver, PSC, PBC, and ARLD on immunohistochemistry (A-B, representative of n=5 patients per condition). Multiplex immunofluorescence showing expression of JAG1, NOTCH2, LSMA (ACTA2), and CK19, in human liver tissue (C, representative of n=5 patients per condition).

## Discussion

Since the widespread use of next generation sequencing, it has become clear that onset and severity of acquired liver diseases (e.g. steatotic liver disease, viral hepatitis) are influenced by germline variation^30,41^. However, these were generally thought to be distinct from the pathways perturbed by rare, loss-of-function variants that lead to early onset severe phenotypes. Building on work done by others^42^, we have demonstrated that common variation in such genes impacts the onset and severity of liver disease in adults. Understanding the pathways implicated in rare, paediatric liver disease may lead to novel therapeutic targets^43^.

It has long been recognised that there is wide phenotypic variation in apparently monogenic disorders^2^. For example, it is not uncommon to have a child with Alagille syndrome affected with profound cholestasis whilst their parent is asymptomatic, despite both harbouring the same variant. Therefore, such pathways are subject to gene-environment interactions, as are other genes where common variants affect liver phenotypes (e.g. *PNPLA3*)^44^. For example, obesity and insulin resistance leading to steatosis promote liver injury in the context of variants that perturb hepatocyte metabolic function (e.g. *LIPA*, where common variants are associated with ALT in this study).

JAG1-NOTCH signalling is a key component of the ductular reaction, being specifically expressed in injured bile ducts, as well as playing a role in liver regeneration in mice^40^. Our observations are consistent with previous murine models showing the role of JAG1-NOTCH in the ductular reaction in steatohepatitis^45,46^. Collectively, the JAG1-NOTCH signalling is one example of how a pathway typically thought of to cause exclusively rare liver disease in early childhood is involved in adult liver disorders.

The concept of ’monogenic’ disease has been progressively eroded across liver disease^9,42^ and other fields^47^. For example, variation in genes that cause monogenic developmental disorders cause more mild phenotypes of the same conditions in the general population^48^. Similarly, for coronary artery disease, genome-wide polygenic risk scores have shown that individuals in the top decile of polygenic risk have a disease incidence comparable to that of ’monogenic’ familial hypercholesterolaemia^49^. This study has illustrated that a similar principle applies for liver disease: the phenotype that affects patients is a combination of polygenic risk (from both rare and common variants) plus environmental factors, even for conditions typically thought of as exclusively genetic or acquired.

These data help in prioritisation of genes for potential therapy and insights into disease mechanisms by integration of these genetic associations with known biological function. For example, *HNF1B* is a promising candidate to investigate the mechanism biliary disorders (e.g. PSC): loss of function variants in *HNF1B* lead to ductal plate malformations, variants in this gene are associated metrics of biliary injury, and its expression is enriched in cholangiocytes^50^. Similarly, *GALK* may be a candidate for steatotic liver disease, given that loss of function variants cause parenchymal injury, variants in *GALK* are associated with ALT, and it is expressed in hepatocytes^50^. GALK inhibitors are already in development for the treatment of classical galactosemia due to the effect of GALK in generating toxic galactose-1-phosphate^51^.

Identification of such effects requires large studies to demonstrate small effect sizes. Overall the effect sizes appear to be about a third compared to common, well-established genome-wide significant variants (e.g. *PNPLA3*). Therefore, it is hard to quantify how these observations influence the phenotype of common liver diseases at an individual patient-level. We observed several variants to reduce the risk of liver diseases, the mechanism of which has not been identified at this time Therefore to elucidate gene- and variant-specific mechanisms will require careful functional studies with cellular stressors.

## Conclusion

Most genes that cause severe monogenic paediatric liver disease are associated with liver injury in adults. The JAG1-NOTCH pathway is a key component of the ductular reaction that is affected by common variation and associated with disease severity in cirrhosis. These findings challenge the notion of ’monogenic’ liver disease and underscore the potential to pharmacologically target the pathways implicated in rare liver diseases for treatment of common diseases in adults.

## Supporting information

SupFig 1

## Data Availability

All raw data is publicly available and code used is available at: https://github.com/jmann01/MonogenicLiver.

## Funding

AS, SPD, YHO, and JPM: NIHR BRC Birmingham.

JPM is supported by grants from Birmingham Health Partners, NIHR ACL (CL-2022-09-005), Royal Society (RG\R1\241312), BSPGHAN-Guts (BSPGHAN2023_01), ESPGHAN, Royal

College Physicians Dame Sheila Sherlock Bursary, and Little Princess Trust (CCLGA 2024 10 Mann).

## Conflicts of interest

KZ is an employee of Gilead Sciences Inc. JX was an employee of Gilead Sciences Inc. at the time the research was conducted.

### Acknowledgements

We thank Gabriel Cuella Partida for his advice on statistical genetics. We also thank the Birmingham NIHR Biomedical Research Centre, Centre for Liver and Gastrointestinal Research, and University of Birmingham Imaging Facility for their support.

## Contributions

CRediT: Mushi J: Data curation, Formal Analysis, Visualization, Writing – original draft, Writing – review & editing; Sharma PJ: Data curation, Formal Analysis, Visualization, Writing – original draft, Writing – review & editing; Schofield A: Data curation, Formal Analysis; Chen VL: Data curation, Formal Analysis, Writing – review & editing; Cordell HJ: Data curation, Formal Analysis, Writing – review & editing; Davies SP: Data curation, Formal Analysis, Writing – review & editing; Gupte GL: Data curation, Formal Analysis, Writing – review & editing; Hirschfield GM: Data curation, Formal Analysis, Writing – review & editing; Jeyaraj R: Data curation, Formal Analysis, Writing – review & editing; Jones DE: Data curation, Formal Analysis, Writing – review & editing; Mells GF: Data curation, Formal Analysis, Writing – review & editing; Oo YH: Data curation, Formal Analysis, Writing – review & editing; Sandford RN: Data curation, Formal Analysis, Writing – review & editing; Siminovitch KA: Data curation, Formal Analysis, Writing – review & editing; Xu J: Data curation, Formal Analysis, Funding acquisition, Investigation, Writing – review & editing; Zhu K: Data curation, Formal Analysis, Funding acquisition, Investigation, Writing – review & editing; Trauner M: Data curation, Formal Analysis, Funding acquisition, Investigation, Methodology, Writing – review & editing; Mann JP: Data curation, Formal Analysis, Funding acquisition, Investigation, Methodology, Project administration, Resources, Supervision, Visualization, Writing – original draft, Writing – review & editing

## Supplementary data

SupTab 1. Genes included in this study and their respective disorders with Online Mendelian Inheritance in Man (OMIM) identifiers.

SupTab 2. HuGE scores and evidence levels for all genes and traits included in this study.

SupTab 3. GeneBass burden testing (pLoF, missense, and synonymous) for all genes and traits included in this study.

SupTab 4. Summary of all variant-trait associations identified in this study. Variants are annotated with: predicted protein functional effect (from Ensembl’s Variant Effect Predictor) and liver tissue eQTL data from GTEx.

SupTab 5. Results from colocalization analyses using data from Sakaue et al. for plots displayed in Figure 2B. Posterior probability results: PP.H0.abf = neither trait has a causal variant in the region; PP.H1.abf = only trait 1 has a causal variant; PP.H2.abf = only trait 2 has a causal variant; PP.H3.abf = both traits have a significant association but the causal variants are different; and PP.H4.abf = both traits have a significant association share the same causal variant. PP.H4.abf >0.8 was described as “Strong” evidence of colocalization.

SupTab 6. Summary of all results from validation of variants in external cohorts.

SupTab 7. Variant-trait associations present in the All of Us dataset. Only significant associations are shown (using Bonferroni adjusted p-value cut-off of 0.05/35 = .0014).

SupTab 8. Results from multivariable logistic regression in the All of Us dataset for the presence of liver disease against monogenic probability risk score (PRS), adjusted for age and sex. PRS was calculated as the sum of alternate alleles for all 35 variants present in the All of Us dataset.

SupTab 9. Results from ANOVA and F0 vs F4 from GSE135251: transcriptomic profiling across the spectrum of MASLD.

SupFig 1. Hepatic expression of JAG1, NOTCH1, HEYL, and HES1 in patients with PSC grouped by Ishak fibrosis stage, collagen deposition, ELF score, and ALP level. Whiskers extend to the minimum and maximum values of the data set. p values are based on the Kruskal–Wallis test. ALP, alkaline phosphatase; ELF, enhanced liver fibrosis; TPM, transcripts per million.

