## Supplementary figures and images for "Genes that cause severe liver disease in children also influence risk and severity of common liver conditions in adults"

### SupFig 1

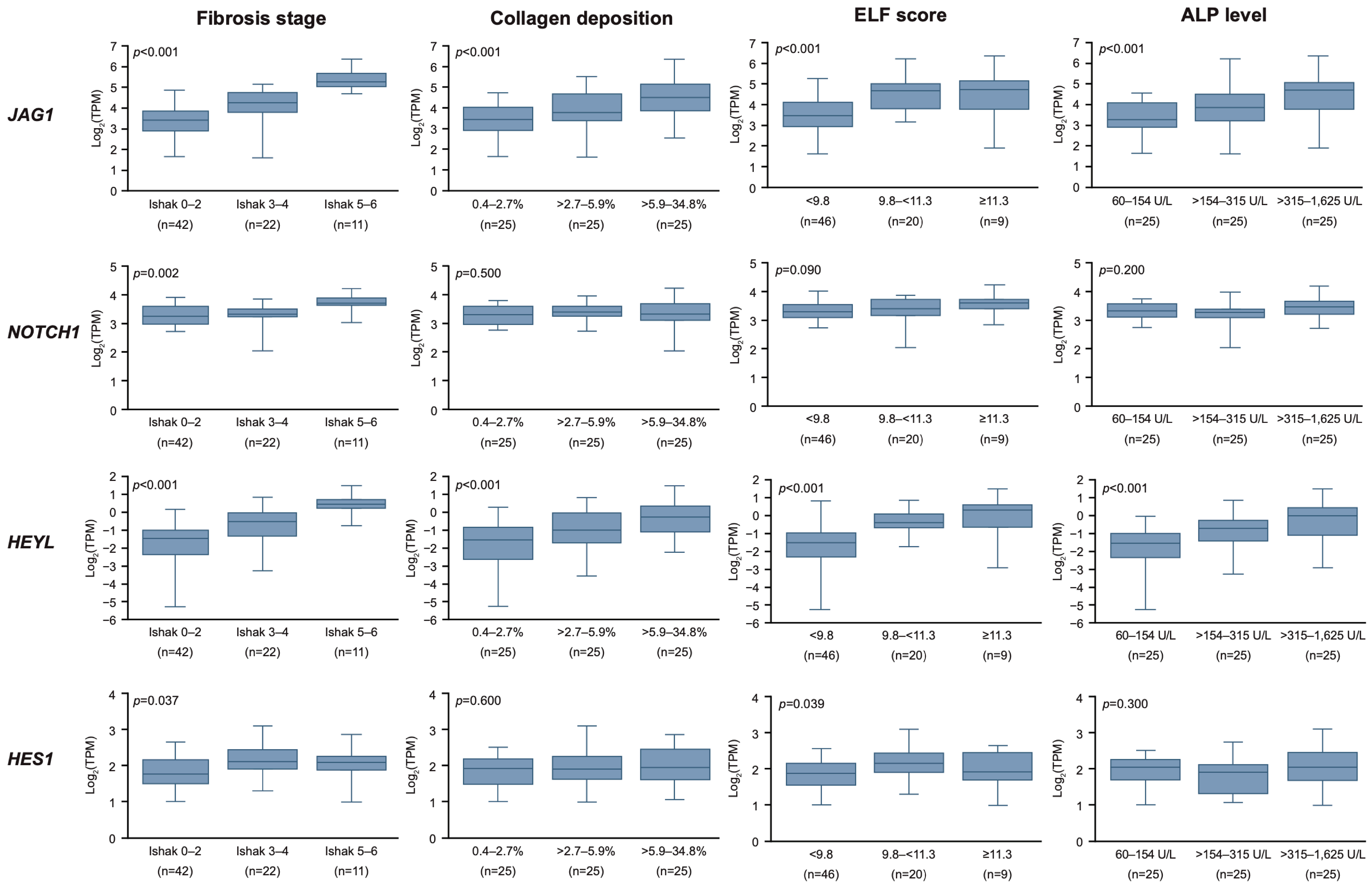
